# Proximity to livestock farms and COVID-19 in the Netherlands, 2020-2021

**DOI:** 10.1101/2022.07.05.22277177

**Authors:** Lenny Hogerwerf, Pim M. Post, Ben Bom, Wim van der Hoek, Jan van de Kassteele, Annette M. Stemerding, Wilco de Vries, Danny Houthuijs

**Affiliations:** National Institute for Public Health and the Environment (RIVM), Bilthoven, the Netherlands; Department of Natural Resources, University of Twente, the Netherlands; Deventer Hospital, Deventer, the Netherlands

## Abstract

**Objectives:** In the Netherlands, during the first phase of the COVID-19 epidemic, the hotspot of COVID-19 overlapped with the country’s main livestock area, while in subsequent phases this distinct spatial pattern disappeared. Previous studies show that living near livestock farms influence human respiratory health and immunological responses. This study aimed to explore whether proximity to livestock was associated with SARS-CoV-2 infection.

**Methods:** The associations between residential (6-digit postal-code) distance to the nearest livestock farm and individuals’ SARS-CoV-2 status was studied in multilevel logistic regression models, comparing individuals notified with a positive SARS-CoV-2 test to the general population in the Netherlands. Data included all reported Dutch laboratory-confirmed patients with disease onset before 1 January 2022. Individuals living in strongly urbanised areas and border areas were excluded. Models were adjusted for individuals’ age categories, the social status of the postal code area, particulate matter (PM_10_)-and nitrogen dioxide (NO_2_)-concentrations. We analysed data for the entire period and population as well as separately for eight time periods (Jan-Mar, Apr-Jun, Jul-Sep and Oct-Dec in 2020 and 2021), four geographic areas of the Netherlands (north, east, west and south), and for five age categories (0-14, 15-24, 25-44, 45-64 and > 65 years).

**Results:** Over the period 2020-2021, individuals’ SARS-CoV-2 status was associated with living closer to livestock farms. This association increased from an Odds Ratio (OR) of 1.01 (95% Confidence Interval [CI] 1.01-1.02) for patients living at a distance of 751-1000 m to a farm to an OR of 1.04 (95% CI 1.04-1.04), 1.07 (95% CI 1.06-1.07) and 1.11 (95% CI 1.10-1.12) for patients living in the more proximate 501-750 m, 251-500m and 0-250 m zones around farms, all relative to patients living further than 1000 m around farms. This association was observed in three out of four quarters of the year in both 2020 and 2021, and in all studied geographic areas and age groups.

**Conclusions:** In this exploratory study with individual SARS-CoV-2 notification data and high-resolution spatial data associations were found between living near livestock farms and individuals’ SARS-CoV-2 status in the Netherlands. Verification of the results in other countries is warranted, as well as investigations into possible underlying mechanisms.

## Introduction

The first case of COVID-19 in the Netherlands was reported on 27 February, 2020. This patient was living in the province of Noord-Brabant in the south of the Netherlands. In the subsequent weeks, it became apparent that COVID-19 incidence remained elevated and largely concentrated in the eastern part of Noord-Brabant and the north of the province of Limburg in the southeast of the Netherlands during the first months of the epidemic (Figure 1 panel A).

**Figure 1.**
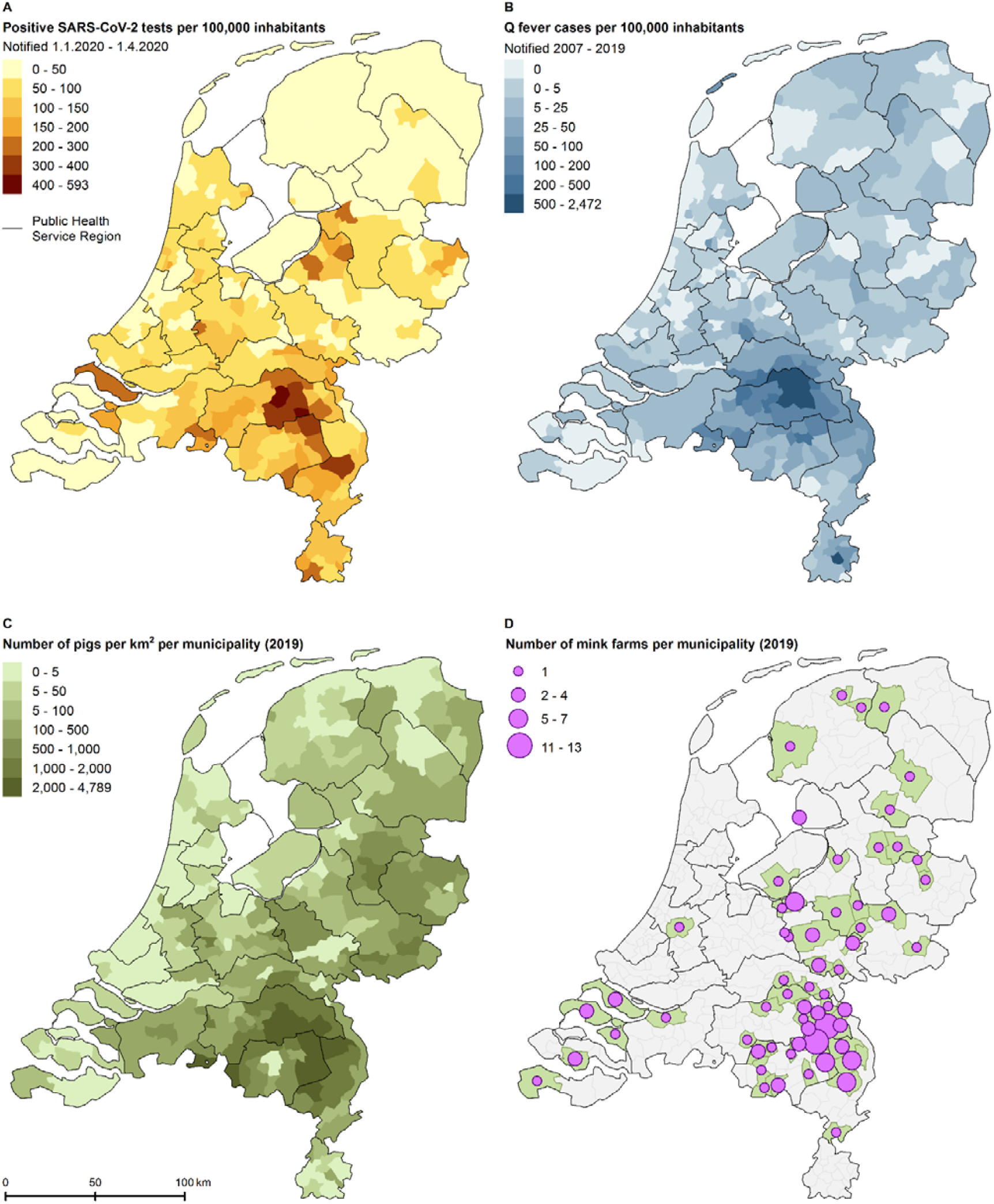
Panel plot with maps of the Netherlands, illustrating the spatial patterns that triggered societal discussion about the relation between COVID-19 and livestock farming (panel A-D). A: Notified COVID-19 deaths per municipality between 31 January and 5 April 2020. B: Notified Q fever cases per municipality during 2007 – 2009. C: Number of pigs per km^2^ per municipality in 2019. D: Number of mink farms per municipality in 2019.

The early COVID-19 hotspots were in part explained by the multiple introduction events involving infected persons who had returned from February holidays spent in northern Italy and Austria, with increased spread by intensive local Carnival celebrations at the end of February. East Noord-Brabant and northern Limburg however, are regions with a high density of livestock farms, including pig, poultry, mink, cattle, goats and others (Figure 1 panel C,D, Figure S1), and had also been the epicentre of the large goat-related Q fever pneumonia epidemic, running from 2007 to 2010 (panel B). This triggered societal discussion about a possible relation between COVID-19 and livestock farming.

Epidemiological studies conducted after the Q fever epidemic learnt that residential proximity to livestock farms was associated with various health effects (Smit and Heederik, 2017), including increased symptoms in patients with chronic obstructive pulmonary disease (COPD) (Borlée et al., 2015; van Dijk et al., 2016; van Kersen et al., 2020) but a lower prevalence of asthma and COPD (Borlée et al., 2015; de Rooij et al., 2019; Post et al., 2021; Smit et al., 2014), a reduced lung function when living close to many farms (Borlée et al., 2017), an increased risk of pneumonia close to goat farms (Freidl et al., 2017; Kalkowska et al., 2018; Klous et al., 2018; Post et al., 2019), higher mortality from respiratory diseases close to pig farms, and indications of higher pneumonia mortality close to mink farms (Simões et al., 2022).

In this exploratory study, we investigated whether individuals’ SARS-CoV-2 status was associated with residential proximity to livestock farms, and whether results were consistent across geographic regions, time periods, and age categories.

## Materials and methods

### Study population

Our study was based on data on the Dutch population on 1 January 2019 and notified SARS-CoV-2 infected patients with an estimated symptom onset during 2020 – 2021.

#### Patients

Laboratory-confirmed SARS-CoV-2 is mandatory notifiable in the Netherlands. We included all notified patients with disease onset, a positive test result, or a notification date before 1 January 2022 in this study. For this, on 4 February 2022, data were extracted from the national database at the National Institute for Public Health and the Environment, to which all 25 Public Health Services in the Netherlands report the laboratory-confirmed SARS-CoV-2 cases. Patient data included age, self-reported date of disease symptoms onset and the six-digit postal code of the residential address. Six-digit postal-code areas comprised on average about eighteen residential addresses. In case date of disease onset was not registered, the date of the laboratory test result or else the date of notification to the Public Health Service was used as proxy. Patients with disease onset in 2022 or for whom the database lacked information on postal code or age were excluded.

#### General population

To be able to compare patients to the general population, a synthetic study population was constructed with a spatially-representative age distribution. Population statistics were retrieved from Statistics Netherlands, which annually provides publicly available statistical data on municipalities, districts, and neighbourhoods (Prins, 2000). A neighbourhood is a part of a municipality that is seen as homogeneously based on historical or urban planning characteristics; it is the smallest area for which the age distribution is publicly available. The Netherlands had 13,379 neighbourhoods on 1 January 2019 with an average population of about 1,300 inhabitants. The digital population on 1 January 2019 of 436,748 six-digit postal-code areas from all 355 municipalities of the Netherlands was apportioned by age distribution into five classes (0-14, 15-24, 25-44, 45-64 years, and 65 years and older) according to Hamilton’s method (Kohler and Zeh, 2012) based on the information available per neighbourhood. This resulted in synthetic age-group-specific population numbers per six-digit postal code. To combine the patient populations with the total synthetic population, the number of notified patients per combination of six-digit postal code and age group were removed from the total synthetic number of inhabitants to obtain the number of non-patients per combination of six-digit postal code and age group. In case this procedure led to a negative number of non-patients for the combination, the number of non-patients was set to zero.

#### Excluded postal code areas

The populations living in very strongly urbanised areas (≥2,500 addresses per km^2^) were excluded from the statistical analyses because the population in these areas tends not to live in proximity to livestock farms, and to exclude risk factors associated with living in these areas. The populations living in six-digit postal codes known to include a nursing home were excluded because of possible data quality issues for nursing homes. Locations of livestock farms in Belgium and Germany were not known, therefore, persons living in six-digit postal codes within two km of the border with Belgium or Germany were excluded.

#### Distance to livestock farms

The distances of the centroid of address locations per six-digit postal-code area to the nearest livestock farm were calculated with ArcGIS 10.6 (ESRI [Environmental Systems Research Institute], 2011), based on information about locations of farms according to the national agricultural census of 1 April 2018 (re: horses, pigs, and poultry); the identification and registration data of 1 July 2019 (re: cattle, goats, and sheep); a list of active farm locations from the Netherlands Food and Consumer Product Safety Authority from 15 June 2020 (re: mink and rabbits). Only farms with a minimum number of animals were included: cattle farms (at least 5 animals), pig farms (≥25), poultry farms (≥200), goat farms (≥50), sheep farms (≥50), horse farms (≥20), rabbit farms (≥200), and mink farms (≥200). The distances to livestock farms of any type were based on the minimum Euclidean distance of the six-digit postal-code centroid to the closest farms of any type. Based on the distances to various types of livestock farms, we defined exposure bands of 0-250, 251-500, 501-750 and 751-1000 meter, with > 1000 meter as reference category, for livestock farms of any type.

### Contextual variables

#### Air pollution

Since ambient air pollution was indicated as a possible risk factor for COVID-19,we included air pollutants with the known largest health impact in the Netherland: particulate matter (PM_10_) and nitrogen dioxide (NO_2_). The modelled annual concentration of PM_10_ and NO_2_ for 2019 was assessed for each six-digit postal code by linking maps yielding 1 × 1 km^2^ grids of the concentrations to all residential addresses in the Netherlands on 1 January 2019, then averaging the concentrations per six-digit postal-code area. These were included as continuous variables in the statistical models. The PM_10_ and NO_2_ concentrations were calculated with the Operational Priority Substances (OPS) dispersion model, which takes into account dispersion, transport, chemical conversion, deposition, and the meteorological conditions in 2019 (Sauter et al., 2018; Van Jaarsveld and De Leeuw, 1993). Source data for this OPS model were the 2018 emissions reported to the Netherlands Pollutant Release and Transfer Register (Wever et al., 2020) and emissions from neighbouring countries (EMEP/CEIP, 2020). NO_2_ levels were derived from the modelled NO_x_ concentration using an empirical relationship between measured NO_x_ and NO_2_ concentrations (van de Kassteele and Velders, 2006; Velders et al., 2014). The concentrations of particulate matter and NO_2_ were calibrated against results from Air Quality Monitoring Networks at 35-45 rural and urban background locations in the Netherlands (number depends on the contaminant). The modelled ambient concentrations represented the average of spatial background concentrations with a resolution of about 1 × 1 km^2^ (Velders et al., 2020).

#### Social status

To adjust for contextual confounding due to social status, we used a social status score at the four-digit postal-code level (on average, 1,987 residential addresses), derived most recently for 2017 by the Netherlands Institute for Social Research. This score is based on income level, unemployment rate, and education level (Knol, 1998), and was standardised with an average of zero and a standard deviation of one. A low social status for a postal code was indicated by a low score. The social status scores were applied to all six-digit postal-codes in the four-digit postal-code area (on average, 109 six-digit postal code areas per four-digit area) to be included as continuous variable in the statistical models.

#### Urbanisation

The degrees of urbanisation of the six-digit postal-code areas were based on statistical data for 2018 from Statistics Netherlands pertaining to 500 × 500-meter squares. This indicator was based on the average address density within a radius of one kilometre divided into five categories: very strongly urbanised (≥2,500 addresses per km^2^), highly urbanised (1,500-2,499 addresses per km^2^), moderately urbanised (1,000-1,499 addresses per km^2^), low-urbanised (500-999 addresses per km^2^) to non-urban (<500 addresses per km^2^).

### Statistical analyses

#### Models

To estimate associations (odds ratio OR and 95% confidence interval CI) between SARS-CoV-2 status and distance to livestock farms, we applied logistic regression models using a random effect for the regional catchment areas of the 25 Public Health Services in the Netherlands. Included covariates were age category, social status score, and air pollution (PM_10_ and NO_2_).

Data management was carried out in Stata version 16 (StataCorp, 2019) and in R version 4.0.1 (R Core Team, 2022). Statistical analyses were performed with R, using glmer function of the lme4 package for the multilevel regression analyses (Bates et al., 2017).

#### Stratifications by epidemic phases, geographic regions and age categories

In addition to analyses of cumulative positive SARS-Cov-2 tests for the entire study period in all studied postal code areas, subsets of the data were explored. This was done to assess whether obtained results were robust over space, time, age groups, and urbanicity levels, and not mainly driven by differences in testing behaviour or virus exposure due to e.g. changing testing policies, triage in hospitals, virus variants, immunity build-up, and the levels of community transmission at the time when social distancing measures were implemented.

Separate analyses were performed for each of eight phases: four quarters of each studied year (January-March, April-June, July-September and October-December in 2020 and in 2021). Each individual notified with a positive SARS-CoV-2 test was assigned to a phase based on the date of symptom onset. In the prior phases they were treated as population control while in subsequent phases they were treated as immune and therefore excluded.

And separate analyses were performed for five age classes (0-14, 15-24, 25-44, 45-64 years, and 65 years and older), the four geographic regions in The Netherlands (North, East, West and South), according to the NUTS (Nomenclature of territorial units for statistics) level 1 classification in the European Union as depicted in Figure S2.

#### Sensitivity analyses

To assess as sensitivity analysis whether inclusion of the covariates affected the outcomes, we performed the analyses without the age categories, without the social score, without the air pollution variables, and with only age as covariate. To assess if nonlinear relations between SARS-CoV-2 status and covariates may have affected the outcomes, we performed three sensitivity analyses using categories based on quintiles for social scores, PM_10_ and NO_2_ concentrations. Two additional sensitivity analyses were performed to evaluate whether the removal of border areas and areas with a nursing home influenced the results. Separate analyses were performed for the four degrees of urbanisation (highly urbanised, moderately urbanised, low-urbanised, and non-urban) and for the the very strongly urbanised areas that had been excluded from the main study population.

Finally, to assess exploratory whether obtained results were driven by particular livestock species, we performed analyses separately per farm type: cattle farms (at least 5 animals), pig farms (≥25), poultry farms (≥200), goat farms (≥50), sheep farms (≥50), horse farms (≥20), rabbit farms (≥200), and mink farms (≥200). For livestock farms of any type and for each farm type, we excluded populations living over 10 km. Next, we defined quintile exposure bands for each farm type, resulting in an equal distribution of the study populations across the distance bands (Table S1). Case data for the entire study period were used, except for analyses on mink farms. Due to SARS-CoV-2 infections in mink farms that were first detected in April 2020, all mink were culled between June and December 2020. Therefore, the analyses on mink farms were performed separately for the first quarter of 2020 (before culling) and for the remaining months of 2020, before mink farming was prohibited in January 2021.

### Privacy

Dutch Civil Code allows the use of health records for statistics or research in the field of public health under strict conditions. All data management and statistical analyses were carried out within the National Institute for Public Health and the Environment. COVID-19 is listed as a notifiable disease and SARS-CoV-2 is listed as a notifiable causative agent in law. No informed consent from patients nor approval by a medical ethics committee is obligatory for registry-based health studies of this type.

## Results

### Population characteristics

As of 1 January 2019, the total Dutch population consisted of 17,278,309 inhabitants. The merger of population data with the individual patient data and exclusion of postal code areas with a missing social status and with nursing home presence, the very strongly urbanised areas and the 2 km border zone with Belgium or Germany led to a synthetic study population of 12,628,244 individuals with full data on exposure at residential address and potential confounders. The majority of exclusions was due to living in a very strongly urbanised area (87%), followed by border areas (10%).

The cumulative number of notified individuals with a positive SARS-CoV-2 tests on the date of database assessment (4 Feb 2022) with disease onset in 2020 or 2021 was 3,190,258, of which 3,121,352 were primo infections. Among these, 3,089,123 had available data on age and a valid postal code and could be merged with the population data. After exclusion of areas with a missing social score, presence of a nursing home, very strongly urbanised areas and the 2 km border zone with Belgium or Germany, 2,223,692 cases remained and were included in the study.

Figure S2 provides a map of the included postal code areas. Figure 2 provides an epicurve of the notified positive SARS-CoV-2 tests in the study population, and Figure S3 shows how these cases are distributed spatially during the eight phases that we distinguished in this study.

**Figure 2.**
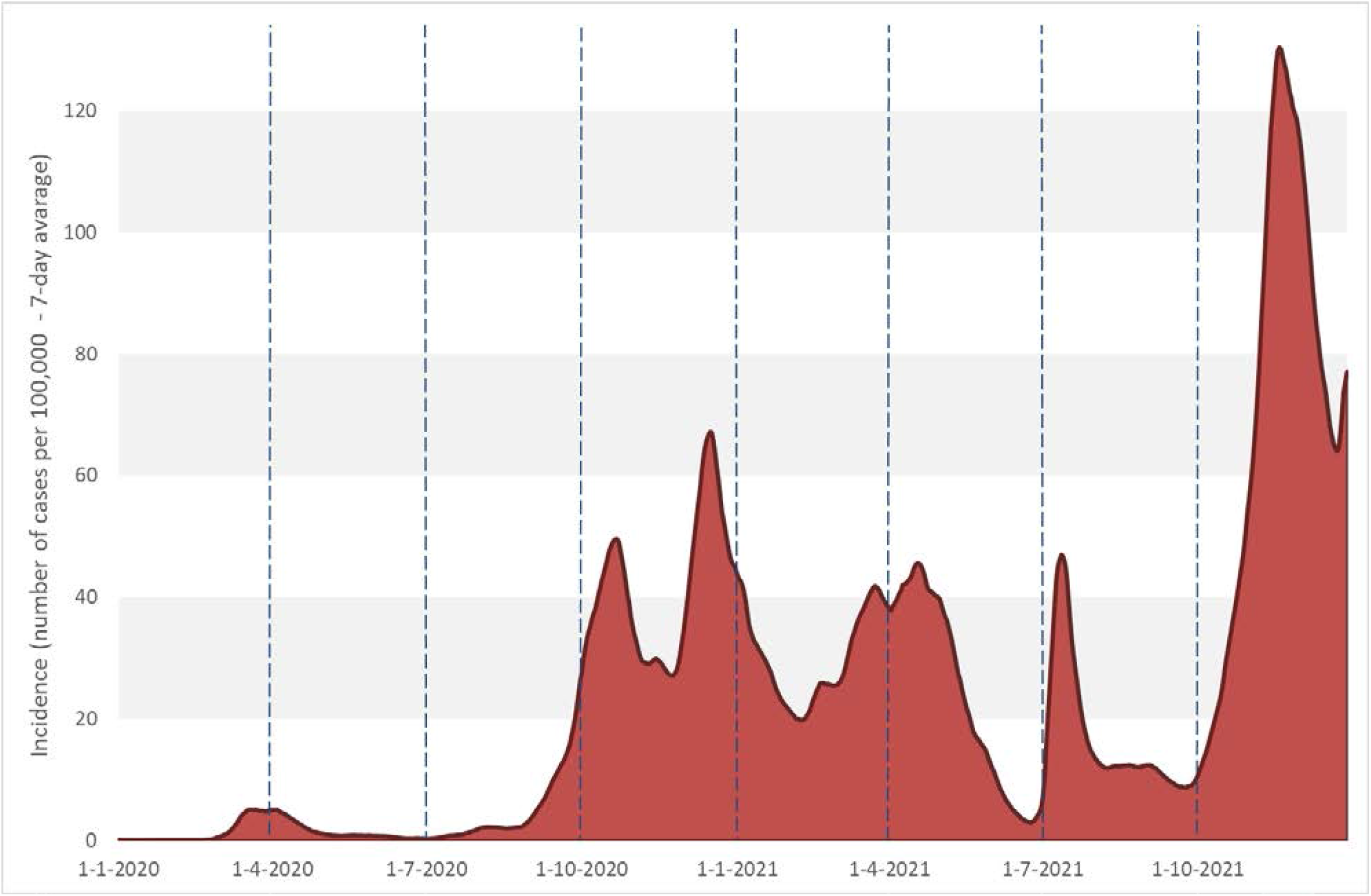
Rolling 7-day average incidence of reported positive SARS-Cov-2 tests by date of disease onset in 2020-2021, shown for the study population.

The characteristics of the study population, including the distributions of the distances to the nearest livestock farms, are depicted in Table 1. Over the period 2020 – 2021, individuals notified with a positive SARS-CoV-2 test were younger than the total study population. Further, notified SARS-CoV-2 cases lived less often in region North, in postal-code areas with on average higher air pollution levels and similar social status scores, and more often in the closer distance bands to livestock in comparison to the total study population. Of the total study population, 38.5% lived within 1 km (Table 1) and all lived within 6.5 km from the closest livestock farm.

**Table 1.** Characteristics of the study population in rural areas in the Netherlands.

| Characteristic | Total population<sup>a</sup> | Individuals notified with a positive SARS-CoV-2 test and symptom onset before 1 January 2022<sup>a,b</sup> |
| --- | --- | --- |
| n | 12.628.244 | 2.223.692 |
| **Age category:** |  |  |
| 0-14 (%) | 16.5 | 14.9 |
| 15-24 (%) | 11.9 | 17.2 |
| 25-44 (%) | 22.6 | 27.9 |
| 45-64 (%) | 29.0 | 28.3 |
| 65 and older (%) | 20.0 | 11.6 |
| **Distance to the nearest livestock farm:** |  |  |
| > 1 km (%) | 38.5 | 38.0 |
| 751-1000 m (%) | 16.3 | 16.1 |
| 501-750 m (%) | 19.1 | 19.2 |
| 251- 500 m (%) | 17.1 | 17.4 |
| 0-250 m (%) | 8.9 | 9.3 |
| **Ambient air pollution:** |  |  |
| Annual average concentration of PM<sub>10</sub> in 2019: |  |  |
| Mean (µg/m<sup>3</sup>) | 17.2 | 17.3 |
| 5-percentile (µg/m<sup>3</sup>) | 14.6 | 14.7 |
| 25-percentile (µg/m<sup>3</sup>) | 16.3 | 16.5 |
| 75-percentile (µg/m<sup>3</sup>) | 18.2 | 18.2 |
| 95-percentile (µg/m<sup>3</sup>) | 18.9 | 19.0 |
| Annual average concentration of NO<sub>2</sub> in 2019: |  |  |
| Mean (µg/m<sup>3</sup>) | 16.1 | 16,3 |
| 5-percentile (µg/m<sup>3</sup>) | 10.2 | 10.6 |
| 25-percentile (µg/m<sup>3</sup>) | 13.6 | 14.0 |
| 75-percentile (µg/m<sup>3</sup>) | 18.2 | 18.4 |
| 95-percentile (µg/m<sup>3</sup>) | 22.5 | 22.7 |
| **Social status postal code:** |  |  |
| Mean score | 0.002 | 0.004 |
| 5-percentile (low) | -2.05 | -2.00 |
| 25-percentile | -0.57 | -0.53 |
| 75-percentile | 0.72 | 0.73 |
| 95-percentile (high) | 1.59 | 1.67 |
| **Region:** |  |  |
| North (%) | 11.6 | 8.8 |
| East (%) | 25.3 | 24.7 |
| West (%) | 40.3 | 41.9 |
| South (%) | 22.7 | 24.6 |
| **Urbanisation degree postal code:** |  |  |
| Highly urbanised (1,500-2,499 addresses per km<sup>2</sup>) (%) | 33.3 | 33.1 |
| Moderately urbanised (1,000-1,499 addresses per km<sup>2</sup>) (%) | 22.6 | 22.3 |
| Low-urbanised (500-999 addresses per km<sup>2</sup>) (%) | 22.5 | 22.8 |
| Non-urban (<500 addresses per km<sup>2</sup>) (%) | 21.7 | 21.8 |

<sup>a</sup> Excluding those living in in a postal code area with a missing social score, with a nursing home, in very strongly urbanised areas, or within two kilometres from the border with Belgium or Germany.
<sup>b</sup> Individuals with a notified positive SARS-CoV-2 test and with estimated symptom onset in 2020 or 2021, excluding those without available data on age and 6-digit postal code area.

### Statistical analyses

#### Distance to livestock farms

People living close to livestock farms had a higher probability of being notified with SARS-CoV-2. Expressed in ORs, there was a trend from an OR of 1.11 (1.10-1.12) in the 0 – 250 m distance band to 1.07 (1.06-1.07) in 251-500 m, to 1.04 (1.04-1.04) in 501-750 m, and to 1.01 (1.01-1.02) in the 751-1000 m distance band compared to > 1,000 metres. Analyses per region, per age group and per phase of the epidemic show similar results, except for the third quarters (July – September) of both studied years (2020 and 2021), the quarters that included periods with the lowest incidences (Table 2).

**Table 2.** Odds ratios (95% confidence interval) for categories of distance to nearest livestock farm (0-250, 251-500, 501-750, 751-1,000 m, and over 1,000 m) for being notified with a positive SARS-CoV-2 test. Results are for the Netherlands and for various subsets (eight quarters, four geographic regions, and five age groups). Excluded from the analyses are residential addresses in areas with a missing social score, with presence of a nursing home, in very strongly urbanised areas, or within 2,000 meters of the border of Germany or Belgium.

| Dataset | Category | n | cases<sup>a</sup> | Distance of residential address to the nearest livestock farm |  |  |  |  |
| --- | --- | --- | --- | --- | --- | --- | --- | --- |
|  |  |  |  | 0 - 250 m | 251-500 m | 501-750 m | 751-1,000 m | > 1,000 m |
|  |  |  |  | OR (95% CI)<sup>b</sup> | OR (95% CI)<sup>b</sup> | OR (95% CI)<sup>b</sup> | OR (95% CI)<sup>b</sup> | ref<sup>b</sup> |
| The Netherlands |  | 12,628,244 | 2,223,692 | 1.11 (1.10-1.12)*** | 1.07 (1.06-1.07)*** | 1.04 (1.04-1.04)*** | 1.01 (1.01-1.02)*** | 1 |
| Subsets |  |  |  |  |  |  |  |  |
| Year 2020 | Jan - Mar | 12,628,244 | 14,252 | 1.08 (1.02-1.16)* | 1.05 (1.00-1.10). | 1.07 (1.02-1.12)** | 1.05 (0.99-1.10). | 1 |
|  | Apr - Jun | 12,613,992 | 16,548 | 1.07 (1.01-1.14)* | 1.03 (0.99-1.08) | 1.04 (1.00-1.09). | 1.02 (0.98-1.07) | 1 |
|  | Jul - Sep | 12,597,444 | 54,983 | 0.93 (0.89-0.96)*** | 0.99 (0.96-1.01) | 0.98 (0.95-1.00)* | 0.97 (0.95-1.00)* | 1 |
|  | Oct - Dec | 12,542,461 | 484,173 | 1.11 (1.09-1.12)*** | 1.07 (1.06-1.08)*** | 1.05 (1.04-1.06)*** | 1.03 (1.02-1.04)*** | 1 |
| Year 2021 | Jan - Mar | 12,058,288 | 342,653 | 1.16 (1.15-1.18)*** | 1.11 (1.09-1.12)*** | 1.07 (1.06-1.08)*** | 1.03 (1.02-1.04)*** | 1 |
|  | Apr - Jun | 11,715,635 | 275,374 | 1.09 (1.08-1.11)*** | 1.04 (1.03-1.06)*** | 1.02 (1.01-1.03)** | 1.00 (0.98-1.01) | 1 |
|  | Jul - Sep | 11,440,261 | 197,605 | 0.99 (0.98-1.01) | 1.01 (1.00-1.02) | 0.99 (0.98-1.00). | 0.99 (0.97-1.00)* | 1 |
|  | Oct - Dec | 11,242,656 | 838,104 | 1.11 (1.10-1.12)*** | 1.06 (1.06-1.07)*** | 1.04 (1.03-1.05)*** | 1.01 (1.00-1.02)* | 1 |
| Region<sup>c</sup> | West | 5,090,497 | 931,404 | 1.10 (1.09-1.11)*** | 1.05 (1.04-1.06)*** | 1.04 (1.03-1.05)*** | 1.01 (1.00-1.01) | 1 |
|  | East | 3,201,038 | 549,839 | 1.17 (1.16-1.18)*** | 1.12 (1.11-1.13)*** | 1.05 (1.04-1.06)*** | 1.02 (1.01-1.03)*** | 1 |
|  | South | 2,871,271 | 546,639 | 1.04 (1.02-1.05)*** | 1.04 (1.03-1.05)*** | 1.01 (1.00-1.02)* | 1.02 (1.01-1.03)*** | 1 |
|  | North | 1,465,438 | 195,810 | 1.11 (1.10-1.13)*** | 1.05 (1.04-1.07)*** | 1.08 (1.06-1.09)*** | 1.04 (1.02-1.05)*** | 1 |
| Age group | 0-14 yo | 2,078,950 | 330,215 | 1.05 (1.04-1.07)*** | 1.07 (1.06-1.08)*** | 1.04 (1.03-1.05)*** | 1.00 (0.99-1.01) | 1 |
|  | 15-24 yo | 1,502,784 | 383,341 | 1.17 (1.16-1.19)*** | 1.07 (1.06-1.08)*** | 1.04 (1.03-1.05)*** | 1.02 (1.01-1.03)*** | 1 |
|  | 25-44 yo | 2,854,256 | 621,282 | 1.10 (1.08-1.11)*** | 1.08 (1.07-1.09)*** | 1.06 (1.05-1.07)*** | 1.02 (1.01-1.03)*** | 1 |
|  | 45-64 yo | 3,667,725 | 629,873 | 1.11 (1.10-1.12)*** | 1.07 (1.06-1.08)*** | 1.04 (1.03-1.05)*** | 1.02 (1.01-1.02)*** | 1 |
|  | ≥ 65 yo | 2,524,529 | 258,981 | 1.13 (1.11-1.15)*** | 1.04 (1.02-1.05)*** | 1.01 (1.00-1.02) | 1.01 (1.00-1.02) | 1 |

. p < 0.1, \* p < 0.05, \*\* p < 0.01, \*\*\* p < 0.001
<sup>a</sup> Individuals with a notified positive SARS-CoV-2 test and with estimated symptom onset before 1 January 2022, excluding those without available data on age and 6-digit postal code area.
<sup>b</sup> Model with 25 regional catchment areas of Public Health Services as random effect adjusted for age category, social status of the four-digit postal-code area, and annual average concentration of PM<sub>10</sub> and NO<sub>2</sub> in 2019 of the six-digit postal-code area
<sup>c</sup> Regions according to NUTS 1

#### Sensitivity analyses

Exclusion of covariates, inclusion of covariates as categorical instead of continuous variables, and the inclusion of border areas or areas with a nursing home did not affect the observed associations (Table S2). Stratification by degree of urbanisation resulted in similar pattens albeit with somewhat lower ORs. Within the very strongly urbanised areas, which were excluded from the study, no associations were seen except one OR slightly below 1 in the 251-500 m distance band.

The use of quintile distance bands for the distance to livestock up to a maximum of 10 km resulted in ORs of 1.09 (1.08-1.09) for the quintile of people closest to farms, to 1.04 (1.03-1.04), 1.01 (1.01-1.02) and 1.00 (1.00-1.01), versus 1 for the reference band (Table 3). Results for cattle, goat, pig, poultry and rabbit farms were similar in size and pattern of the highest ORs for people living the closest to farms with ORs gradually decreasing along the larger distances to farms. For sheep farms, ORs followed a similar pattern but were somewhat lower. Results for mink farms showed a similar pattern, but ORs were different in size for the two studied periods: OR 1.25 (1.15-1.36) for January-March 2020 (prior to the culling of mink) and OR 1.03 (1.02-1.05) for April-December 2020 (during culling until the ban on mink farms) for people living 0-3.55 kilometres from the closest mink farm. Results for horse farms were an exception, with all ORs smaller than 1 and without any pattern along the distances (Table 3).

**Table 3.**
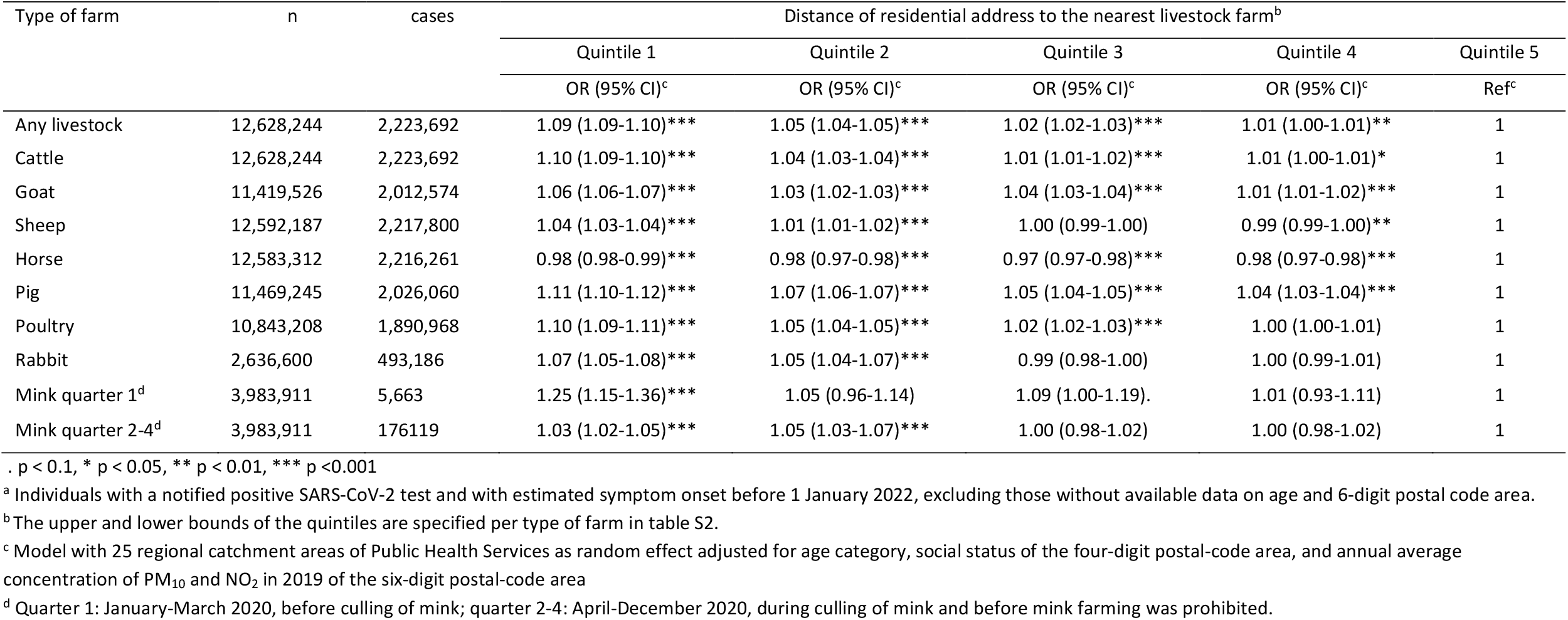
Odds ratios (95% confidence interval) for quintiles of distance to nearest livestock farm and for distance to different farm types for being notified with a positive SARS-CoV-2 test. Results are for the Netherlands and excluded from the analyses are residential addresses with a distance of more than 10 km from the type of farm, in areas with a missing social score, with presence of a nursing home, in very strongly urbanised areas, or within 2,000 meters of the border of Germany or Belgium.

## Discussion

Livestock farm proximity enhanced the probability of individuals to be notified with a positive SARS-CoV-2 test. Results were similar across regions and age groups, and for six out of eight studied time periods. The same association between livestock proximity and SARS-CoV-2 status was observed for the first, second and fourth quarter of 2020 and 2021 but not for the July-September periods, when the incidence was lowest.

Sensitivity analyses per livestock species showed comparable results across farms of any type, cattle, goat, pig, poultry and rabbit farms. For sheep farms, ORs were somewhat lower, possibly related to sheep grazing at locations other than the farm itself. Exposure misclassification is likely to be more limited for other grazing animals. Cattle commonly graze during part of the year, but less than sheep, while most goats remain within the farm. For horses, results were not in line with the other species, with ORs below 1. These ‘farms’ often included horse riding schools, while many other riding school locations were not available to our study, making results for horses hard to interpret. Analyses for mink resulted in higher ORs for the first quarter of 2020, before the culling of mink due to SARS-CoV-2 infections in mink. For the three subsequent periods, during culling and until mink farming in the Netherlands completely stopped by the end of 2020, ORs were reduced. The coherence of results across species suggest that the observed associations are not driven by one particular farm type, despite that in the analyses per species, no adjustments were made for the proximity to other species.

Ambient air pollution has been associated with SARS-CoV-2 incidence in the Netherlands (Andree, 2020; Cole et al., 2020) and in several other countries among others Canada (Stieb et al., 2020), USA (Sidell et al., 2022), Italy (De Angelis et al., 2021) and Germany (Prinz and Richter, 2022). We therefore included ambient PM_10_ and NO_2_ concentrations as covariates. Livestock production is one of the sources that contributes to air pollution, in particular to particulate matter (PM) concentrations, so the associations with distance to livestock farms may have been over adjusted. However, sensitivity analyses without PM_10_ and NO_2_ gave similar results (Table S2) suggesting that overadjustment due to general air pollution is not an issue.

The results of our study are in line with previously reported associations between proximity to livestock farms and various respiratory health outcomes, including lower respiratory infections, where multiple livestock species have been implicated (Freidl et al., 2017; Kalkowska et al., 2018; Klous et al., 2018; Post et al., 2019; Poulsen et al., 2018; Simões et al., 2022; Smit et al., 2012). Hypotheses about underlying biological and physical mechanisms have been proposed. For instance, persons living in livestock areas having an enhanced responsiveness to livestock specific particulate matter (PM) including microbial contaminated PM, or Bio-PM triggering innate immune responses, possibly contributing to airway diseases (Liu et al., 2019; Poole and Romberger, 2012; Sahlander et al., 2012). Possibly, a similar mechanism might enhance the risk of SARS-CoV-2 infection (Diamond and Kanneganti, 2022). But this requires further investigation.

Of the types of farms included in this study, only mink farms have been shown to be infected by humans with SARS-CoV-2 (ECDC, 2020; Enserink, 2020; Oreshkova et al., 2020). Whole genome sequences provided evidence of mink-to-human transmission following genetic evolution in the animals (Oude Munnink et al., 2020). But spill-back of a mink sequence into the community, as occurred in Denmark (Hammer et al., 2021), was not observed in the Netherlands (Oude Munnink et al., 2020) so this route is unlikely to explain our study results for mink farms. In our study, we found similar results for multiple time periods and regions, also in absence of mink. This means that mink were not the main driver of the study outcomes. A similar reasoning applies to the former goat-related Q fever epidemic, of which the main affected area overlapped with the initial COVID-19 hotspot (van Gageldonk-Lafeber et al., 2021; Weehuizen et al., 2022). The associations that we found were not limited to the former Q fever areas.

In recent years, outbreaks of animal coronaviruses have occurred in e.g. pigs (porcine epidemic diarrhoea virus: PEDV) (Dortmans et al., 2018), poultry (de Wit et al., 2021) and horses (equine coronavirus) (Zhao et al., 2019), and many other animal coronaviruses are endemic in the Netherlands and worldwide and present in the environment (Decaro et al., 2020; Khamassi Khbou et al., 2021). One consideration is whether these animal coronaviruses, when inhaled and present on mucus, could result in false positive SARS-CoV-2 tests specifically in people around livestock farms. However, this possible explanation of our study results seems very unlikely as we expect this would have been noticed given ongoing whole genome sequencing activities worldwide.

The main challenge of the study was to avoid possible interference by local, under-the-radar, virus introductions and spread. During the start of the epidemic, SARS-CoV-2 was introduced unevenly frequent across the country, for example by persons returning from February 2020 holidays, and locally amplified by carnival celebrations, but data to reconstruct such spread across networks are sparse. The areas with the highest level of transmission at the moment of the implementation of control measures (lockdown) may have happened to coincide with, in this case, intensive livestock production. When exposure to the virus is unknown, risk estimates for the incidence may be biased and may change as an epidemic progresses (Koopman et al., 1991; Villeneuve and Goldberg, 2022).

While all reported SARS-CoV-2 cases were available to this study, not all infected individuals were tested or reported. Testing policies varied and changed substantially across settings (e.g. for healthcare workers) and over time (restrictive at first, broader later). The effect of testing can be seen in the epicurves in Figure 2, which show that particularly in the first phases of the pandemic testing capacity was very limited. It is unknown if selective underdiagnosis and underreporting affected our results, since it is not known yet if the underreporting is related to the exposures of our interest.

With these two possible issues and dynamics in mind, we performed the analyses for several phases over a period of two years, for several regions, and in various sensitivity analyses. Results for proximity to livestock were consistent, however with ORs turning lower or below 1 during the third quarter of the year.

Our study was able to use individual patient data and the six-digit postal-code of the address of SARS-CoV-2 cases, avoiding some of the inherent limitations of studies that rely on publicly available information at higher aggregation levels (Heederik et al., 2020; Villeneuve and Goldberg, 2020). However, we could not control for individual factors such as comorbidities, household income and education level. In a follow-up study in the Netherlands on air pollution and COVID-19 (Kamerstukken, 2021) (pseudo-anonymised) data at the individual level will be used and more advanced methods to account for the underling human behaviour and transmission will be developed.

## Conclusions

This study suggests that proximity to livestock was associated with individuals’ probability of being notified with a positive SARS-CoV-2 test in 2020-2021 in the Netherlands, but mechanisms are not understood. International replication and verification is needed.

## Supporting information

EQUATOR checklist

## Data Availability

Data produced in the present study are available upon reasonable request to the authors. Restrictions apply to the availability of case-based data.

## Acknowledgements

We would like to thank Lucy Philips for language editing of previous version of this manuscript, Wim Swart for data-management, Frans Corten for a good suggestion, and Erik Lebret for commenting on earlier versions.

## Funding

This study was funded from the regular budget (project V/150207/20/RI), COVID-19 budget (projects D/111001/01/CO and V/190035/22/EB) and the Strategic Program (SPR) (project S/113002/01/IC) of the National Institute for Public Health and the Environment (RIVM), made available by the Ministry of Health, Welfare and Sport of the Netherlands The funding sources had no role in the study design; in the collection, analysis and interpretation of data; in the writing of the manuscript; and in the decision to submit the article for publication. Declarations of interest: none

## Supplementary Material

Supplementary figures and tables

## Notes

### Competing Interest Statement

The authors have declared no competing interest.

### Author Declarations

The Centre for Clinical Expertise at the RIVM assessed the research proposal following the specific conditions as stated in the law for medical research involving human subjects (WMO). In their opinion the research does not fulfill one or both of these conditions and therefore conclude it is exempted for further approval by the ethical research committee. Pathogen surveillance is a legal task of the RIVM (artikel 3 Wet RIVM, article 3 Law RIVM) and is carried out under the responsibility of the Minister of Health, Welfare and Sports (VWS). Article 6c (artikel 6c) of the Public Health Act (Wet Publieke Gezondheid) provides that RIVM may receive pseudonymised data for this task without individual consent of each case.

