## Supplementary figures and tables for "Proximity to livestock farms and COVID-19 in the Netherlands, 2020-2021"

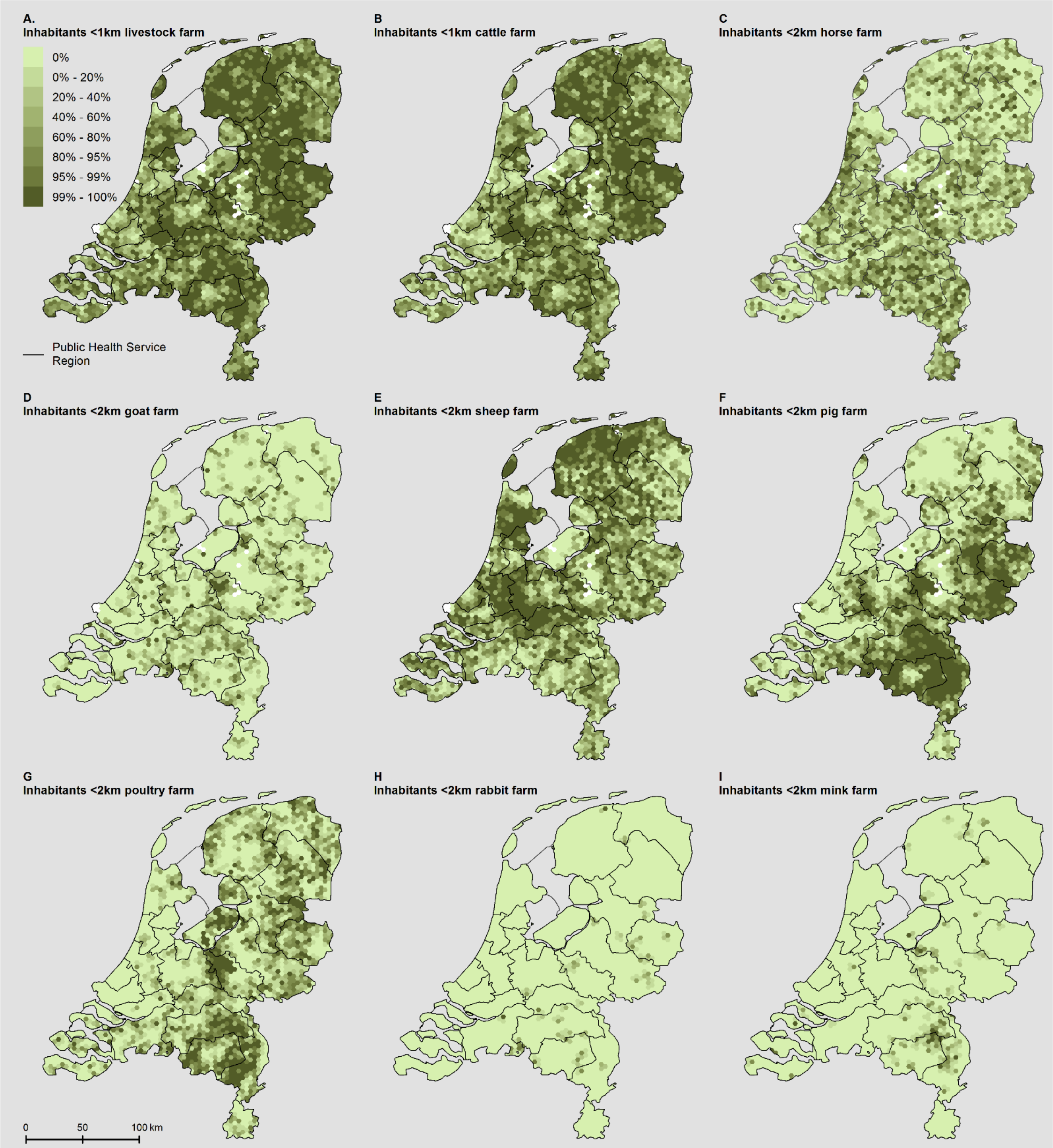


*Figure S1. Nine-Panel plot illustrating the spatial patterns of home address distances to the livestock farm types included in this study.*

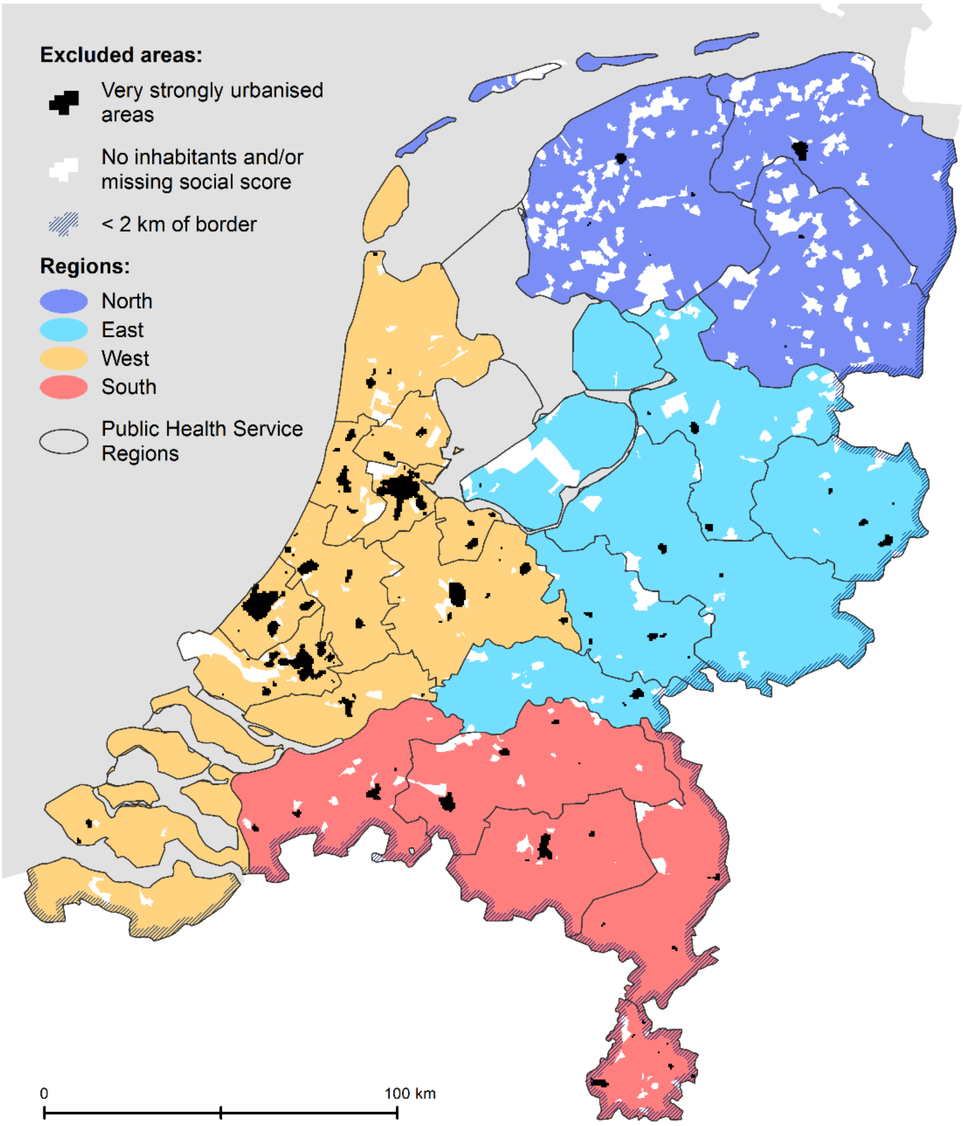


*Figure S2. Map of the Netherlands illustrating the 25 regional catchment areas of Public Health Services, the four regions, and the very strongly urbanised areas (≥2,500 addresses per $km_2$), the area within two kilometres of the border with Belgium or Germany, and areas with no inhabitants and/or a missing social score. Areas with presence of a nursing home were too small in size to be shown.*

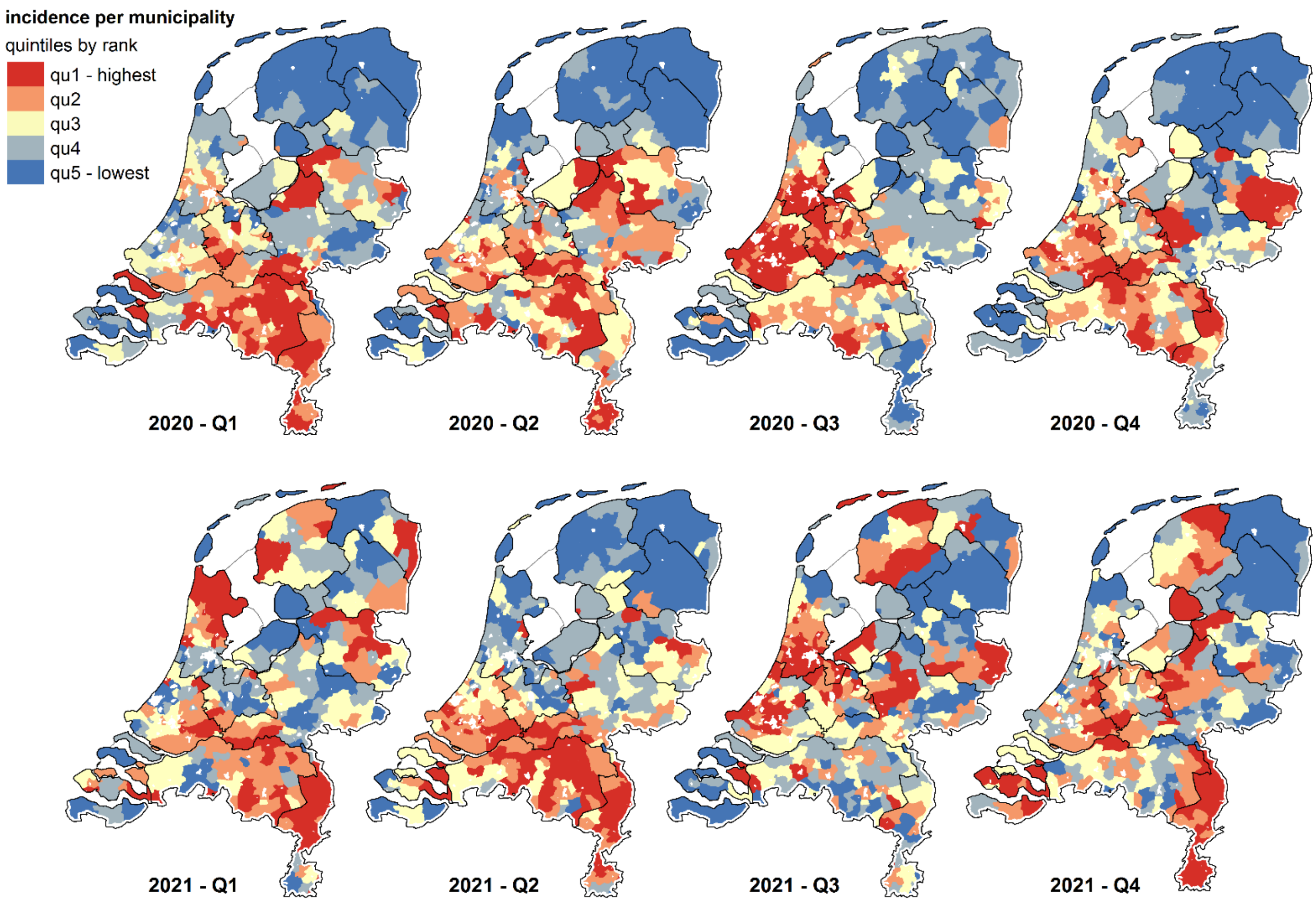


*Figure S3. Eight-Panel plot illustrating notified positive SARS-CoV-2 tests per 100,000 study population per municipality for eight phases of the COVID-19 epidemic (quarters 1 – 4 of 2020 and 2021). For each phase, municipalities were ranked according to the proportion of individuals with a positive test, and categorised in five quintiles (qu1 – qu5). Areas in white indicate the excluded border areas and very strongly urbanised areas. Areas excluded due to a missing social score or presence of a nursing home are not shown.*

Tabel S1. Upper and lower bounds of quintiles of distance of residential address to the nearest livestock farm in kilometres, per type of farm.

| Type of farm | Quintile 1 | Quintile 2 | Quintile 3 | Quintile 4 | Quintile 5 |
|---|---|---|---|---|---|
| Livestock of any type | [0,0.421] | (0.421,0.682] | (0.682,0.975] | (0.975,1.42] | (1.42,6.48] |
| Cattle | [0,0.52] | (0.52,0.836] | (0.836,1.19] | (1.19,1.73] | (1.73,7.44] |
| Goat | [0,2.48] | (2.48,3.6] | (3.6,4.81] | (4.81,6.67] | (6.67,10] |
| Sheep | [0,0.841] | (0.841,1.4] | (1.4,1.98] | (1.98,2.83] | (2.83,10] |
| Horse | [0,1.25] | (1.25,1.86] | (1.86,2.49] | (2.49,3.36] | (3.36,10] |
| Pig | [0,1.28] | (1.28,2.14] | (2.14,3.28] | (3.28,5.3] | (5.3,10] |
| Poultry | [0,1.7] | (1.7,2.76] | (2.76,4.07] | (4.07,6.04] | (6.04,10] |
| Rabbit | [0,4.1] | (4.1,6.01] | (6.01,7.39] | (7.39,8.71] | (4.07,6.04] |
| Mink | [0,3.35] | (3.35,5.11] | (5.11,6.58] | (6.58,8.33] | (8.33,10] |

Table S2. Odds ratios (95% confidence interval) for categories of distance to nearest livestock farm (0-250, 250-500, 500-750, 750-1000 m, and over 1000 m) for being notified with a positive SARS-CoV-2 test. Sensitivity analyses with different combinations of covariates and for different study populations (including borders areas, including postal codes with nursing homes and stratified by urbanisation level).

| Type of analysis | n | cases[a] | Distance of residential address to the nearest livestock farm | | | | |
|---|---|---|---|---|---|---|---|
| | | | 0 - 250 m | 251-500 m | 501-750 m | 751-1000 m | 1-10 km |
| | | | OR (95% CI)[b] | OR (95% CI) [b] | OR (95% CI) [b] | OR (95% CI) [b] | ref [b] |
| Sensitivity analyses[c] | | | | | | | |
| Without age group | 12,628,244 | 2,223,692 | 1.11 (1.10-1.12)*** | 1.07 (1.06-1.07)*** | 1.04 (1.03-1.04)*** | 1.01 (1.01-1.02)*** | 1 |
| Without $PM_{10}$ and $NO_2$ | | | 1.11 (1.11-1.12)*** | 1.07 (1.06-1.07)*** | 1.04 (1.04-1.04)*** | 1.01 (1.01-1.02)*** | 1 |
| Without social score | | | 1.11 (1.10-1.12)*** | 1.07 (1.06-1.07)*** | 1.04 (1.03-1.04)*** | 1.01 (1.01-1.02)*** | 1 |
| Without $PM_{10}$, $NO_2$ and social score | | | 1.11 (1.10-1.12)*** | 1.07 (1.06-1.07)*** | 1.04 (1.04-1.04)*** | 1.01 (1.01-1.02)*** | 1 |
| Social score included as quintiles | | | 1.11 (1.10-1.11)*** | 1.06 (1.06-1.07)*** | 1.04 (1.03-1.04)*** | 1.01 (1.01-1.02)*** | 1 |
| $PM_{10}$ included as quintiles | | | 1.11 (1.10-1.11)*** | 1.07 (1.06-1.07)*** | 1.04 (1.04-1.04)*** | 1.01 (1.01-1.02)*** | 1 |
| $NO_2$ included as quintiles | | | 1.10 (1.10-1.11)*** | 1.06 (1.06-1.07)*** | 1.04 (1.03-1.04)*** | 1.01 (1.01-1.02)*** | 1 |
| Study populations[c] | | | | | | | |
| With border areas | 13,058,100 | 2,300,842 | 1.11 (1.10-1.11)*** | 1.06 (1.06-1.07)*** | 1.04 (1.03-1.04)*** | 1.01 (1.01-1.02)*** | 1 |
| With nursing home postal codes | 12,795,960 | 2,266,936 | 1.11 (1.10-1.11)*** | 1.06 (1.06-1.07)*** | 1.04 (1.03-1.04)*** | 1.01 (1.01-1.02)*** | 1 |
| Urbanisation level: | | | | | | | |
| -Very strongly (≥2,500 addresses/$km^2$) | 3,983,302 | 713,296 | 0.97 (0.94-1.01) | 0.98 (0.96-1.00)* | 1.00 (0.99-1.01) | 1.00 (0.99-1.01) | 1 |
| -Highly (1,500-2,499 addresses/$km^2$) | 4,206,920 | 736,671 | 1.04 (1.02-1.06)*** | 1.01 (1.00-1.02)** | 1.01 (1.00-1.02). | 1.00 (0.99-1.01) | 1 |
| -Moderate (1,000-1,499 addresses/$km^2$) | 2,850,442 | 496,707 | 1.02 (1.00-1.04)* | 1.03 (1.02-1.04)*** | 1.01 (1.00-1.02)** | 1.01 (1.00-1.02). | 1 |
| -Low (500-999 addresses/$km^2$) | 2,836,049 | 506,653 | 1.05 (1.04-1.06)*** | 1.03 (1.02-1.04)*** | 1.03 (1.02-1.04)*** | 1.00 (1.00-1.01) | 1 |
| -Non-urban (<500 addresses/$km^2$) | 2,734,833 | 483,661 | 1.07 (1.06-1.08)*** | 1.04 (1.03-1.05)*** | 1.03 (1.02-1.04)*** | 1.00 (0.99-1.02) | 1 |

. $p < 0.1$, * $p < 0.05$, ** $p < 0.01$, *** $p < 0.001$

[a] Individuals with a notified positive SARS-CoV-2 test and with estimated symptom onset before 1 January 2022, excluding those without available data on age and 6-digit postal code area.

[b] Model with 25 regional catchment areas of Public Health Services as random effect adjusted for age category, social status of the four-digit postal-code area, and annual average concentration of $PM_{10}$ and $NO_2$ in 2019 of the six-digit postal-code area (unless specified otherwise)

[c] Excluding residential addresses in areas with a missing social score, with presence of a nursing home, in very strongly urbanised areas, or within 2,000 meters of the border of Germany or Belgium unless specified otherwise.
